# Cost-effectiveness of psychosocial assessment for individuals who present to hospital following self-harm in England: a model-based retrospective analysis

**DOI:** 10.1101/2021.09.22.21263969

**Authors:** David McDaid, A-La Park, Apostolos Tsiachristas, Fiona Brand, Deborah Casey, Caroline Clements, Galit Geulayov, Nav Kapur, Jennifer Ness, Keith Waters, Keith Hawton

## Abstract

**Aims:** There are substantial costs to health care systems and society associated with self-harm. Moreover, individuals who have presented to hospital following self-harm have a much higher risk of suicide within the following year compared to the general population. National guidance in England recommends psychosocial assessment when presenting to hospital following self-harm but adherence to this guidance is variable. There is some limited evidence suggesting that psychosocial assessment is associated with lower risk of subsequent presentation to hospital for self-harm. The aim of this study was to assess the potential cost-effectiveness of psychosocial assessment for hospital-presenting self-harm in England compared to no assessment.

**Methods:** We constructed a three-state four-cycle Markov model to assess the cost-effectiveness of psychosocial assessment after self-harm compared with no assessment over two years. Data on risk of subsequent self-harm and hospital costs of treating different types of self-harm were drawn from prior analysis of the Multicentre Study of Self-Harm in England, while estimates of the effectiveness of psychosocial assessment on risk of self-harm, quality of life impacts and other costs were supplemented by a literature review. Incremental cost-effectiveness ratios (ICERs) were estimated in terms of cost per Quality Adjusted Life Year (QALY) gained and parameter uncertainty was addressed in univariate and probabilistic sensitivity analyses. Costs were reported in 2020 UK Pounds from the healthcare and societal perspective (that included productivity loss) and a discount rate of 3.5% was applied to future costs and QALYs.

**Results:** The cost per QALY gained from psychosocial assessment was £10,962 (95% uncertainty interval (UI) £15,538 - £9,219) from the NHS perspective, and £9,980 (95% UI £14,538, £6,938) from the societal perspective. Baseline results were generally robust to changes in model assumptions; the relative risk of self-harm after psychosocial assessment would have to be 0.73 or lower for the ICER to be below £20,000. The cost-effectiveness acceptability curve showed that the probability of the ICER to be below a £20,000 threshold was 78%, rising to 91% with a £30,000 threshold.

**Conclusions:** Psychosocial assessment as implemented in the English NHS is likely to be cost-effective. This evidence could support adherence to NICE guidelines. However, further evidence is still needed about the precise impact of psychosocial assessment on self-harm repetition and costs to individuals affected by self-harm and their families beyond immediate hospital stay.

## Introduction

Self-harm, defined as non-fatal intentional self-poisoning or self-injury, irrespective of degree of suicidal intent or other motives (1, 2), is a major health care problem globally. In England it involves over 200,000 hospital presentations per year (3, 4), with substantial costs, many of which are potentially avoidable through better public health and health system actions. In England, hospital costs alone have been estimated to be more than £128 million per year (4). While substantial, these represent just one of the more visible elements of cost; there will be other costs both to health systems and wider society associated with self-harm unrelated to hospital presentation (5). There is therefore an economic, as well as moral imperative, to understand not only what works in preventing self-harm and suicide in the development of policy, but also to determine the relative cost-effectiveness of interventions against alternative investments in the health system to both improve quality of life and reduce premature mortality.

Psychosocial assessment is recommended for all hospital presenting self-harm in England (2, 6). On average around 50-60% of people presenting to accident and emergency (A&E) departments with self-harm receive a full psychosocial assessment, although this proportion varies across sites (7, 8). This clinical procedure, typically carried out by psychiatric liaison staff, includes assessing patients’ problems, mental state, risk factors and needs, and arranging appropriate aftercare. Investment in greater adherence to guidance on use of psychosocial assessment may help reduce risk of repeat self-harm (9), as well as having other potential therapeutic benefits. Psychosocial assessment may therefore be a cost-effective mechanism from a public health perspective but there is relatively little evidence on the economic case for increasing adherence to guidance. This study aimed to model the potential cost-effectiveness of psychosocial assessment for hospital presenting self-harm in England compared to no assessment. The work draws on data on hospital presenting self-harm from the Multicentre Study of Self-harm in England (MSH) (3).

## Methods

### Model type and structure

In the absence of empirical data from trials, health economic modelling studies are widely used to help determine the potential strength of the economic case for action (10). Models bring together evidence on effectiveness, resource use and costs from multiple sources. One of the principal approaches is Markov modelling. It can be used to model uncertain processes over multiple time periods known as cycles and reflect circumstances where individual health and outcomes can fluctuate (11). Markov modelling has been used by public health agencies in England to support local decision makers, for instance to develop their mental health promotion and disorder prevention strategies, including work to prevent bullying in schools and suicide in adults (12).

A three-state Markov model has been constructed. Figure A1 in the supplementary appendix provides an overview of the model’s health states, with a more detailed excerpt shown in appendix Figure A2. All individuals enter the model when initially presenting at a hospital A&E department following a self-harm event. Individuals may be treated in A&E only or may be admitted to hospital for further treatment and observation. Patterns of treatment and length of stay (if admitted) also vary depending on physical severity of self-harm. The model assumes that in each subsequent cycle there are three possible states: no hospital presenting self-harm event, a further A&E presentation following self-harm, and death as a result of suicide.

### Cost-utility analysis

The model runs over two years with each Markov cycle lasting 6 months, comparing receipt of psychosocial assessment following each hospital presenting self-harm event to non-receipt of psychosocial assessment after a self-harm event. The primary model outcome is change in quality of life associated with self-harm. Utility values are assigned to the three health states and QALYs estimated based on time spent in each health state. Mean costs associated with self-harm events in each model cycle were computed for each health state, except for death. All costs and outcomes are reported in 2020 British pounds (£), discounted at a Treasury recommended annual rate of 3.5% after 1 year (i.e. the last 2 six-month cycles). Where necessary, costs have been converted to 2020 prices using UK Office of National Statistics GDP Deflators (13). Our primary analysis is conducted from the publicly funded English National Health Service (NHS) perspective, but we also report results from a societal perspective taking account of productivity losses to patients when in hospital. No other productivity losses, such as impacts on families or the economic impact of premature death through suicide, are included. Cumulative QALYs and total costs over the model’s two-year time horizon have been calculated and incremental costs per QALY gained estimated. A CHEERS (Consolidated Health Economic Evaluation Reporting Standards) recommended reporting checklist for health economic studies (14) is available as Appendix Table A1.

### Sensitivity analysis and uncertainty

Sensitivity analyses have been conducted to assess the robustness of results to underlying input parameters and assumptions. In univariate sensitivity analysis we varied most different individual parameters in the NHS perspective model, one at a time, by up to 20% from their mean values in Table 1. One exception was for the probability of repeat self-harm after psychosocial assessment. In this case in sensitivity analyses we ensured that the lower estimate of effect reflected more conservative values reported in the literature (15–17).

**Table 1:**
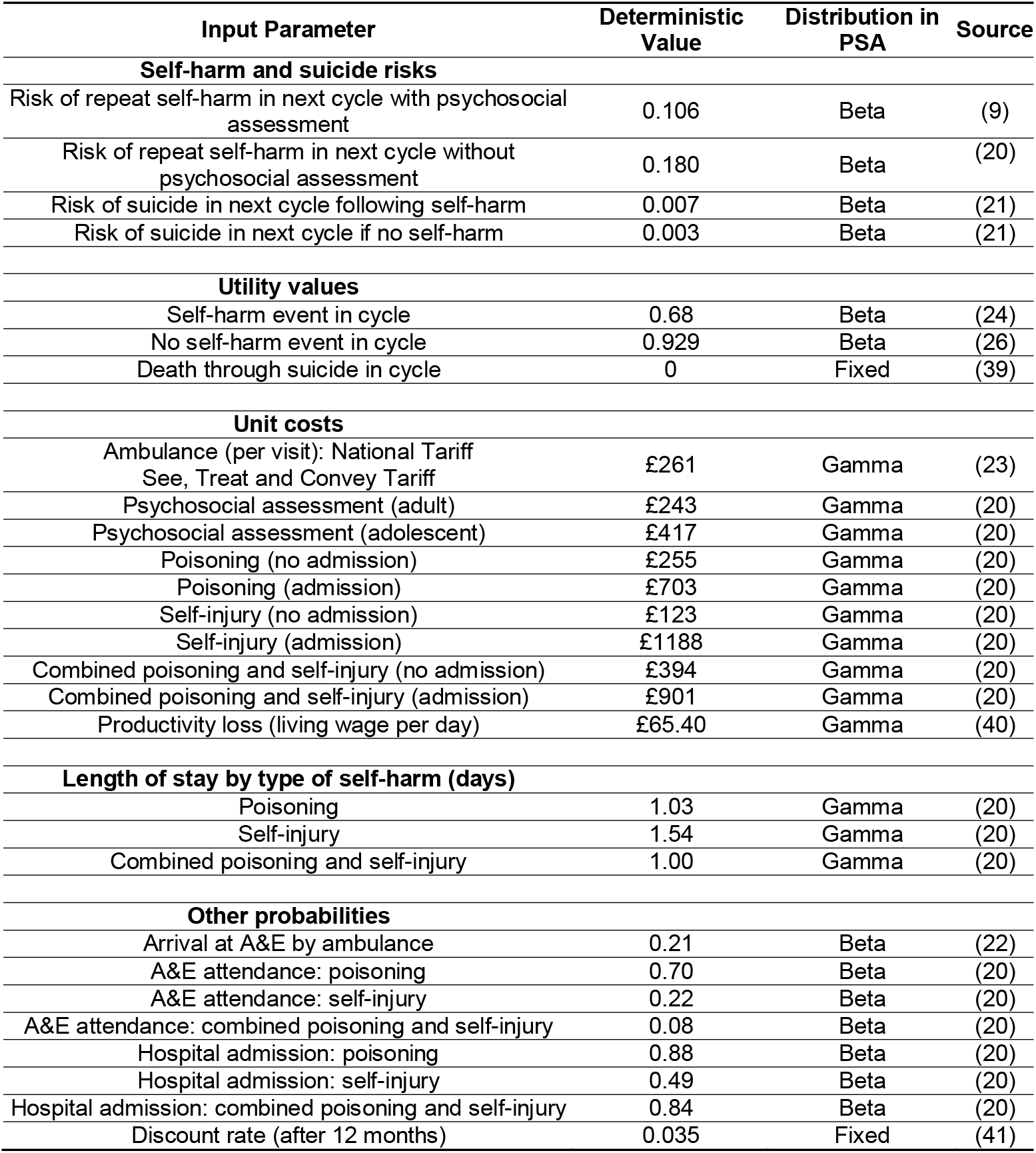
Model parameters.

In addition, we looked at the possible direct impact of psychosocial assessment on suicide. In our baseline model we assumed no risk reduction, given a lack of evidence (18); here we varied possible risk reduction between 0 and 100%. For the societal perspective model we looked at the impact of increasing the value of productivity losses by up to £100 per day for inpatient hospital stays.

We also conducted Monte Carlo simulation, where the uncertainty associated with model variables can be estimated from the parameters’ distribution. This is done by randomly sampling a value for selected parameters from within their distributions simultaneously and then calculating incremental cost-effectiveness. We repeated this exercise 10,000 times. Following best practice, input parameters were assigned beta, gamma or log-normal shaped distributions, as appropriate (19). We visually show results on a cost-effectiveness plane, where all 10,000 combinations of incremental costs and incremental QALYs were plotted. Additionally, histograms showing distribution of net monetary benefits were constructed.

Cost effectiveness acceptability curves, indicating likelihood of the intervention being considered cost-effective at different thresholds of willingness to pay per QALY gained, were generated from both healthcare and broader societal perspectives. All analyses are modelled using TreeAge Pro software (TreeAge Software, LLC, Williamstown, MA, USA).

### Model parameters

Model parameters and assumptions on their distributions in probabilistic sensitivity analysis are shown in Table 1. Hospital presenting self-harm data are taken from the MSH. The MSH systematically collects data, captured via clinicians and clinical records for all hospital presentations for self-harm in Oxford, Derby and Manchester. These areas have diverse populations with varied socio-demographic characteristics, and may therefore, provide a reasonably representative picture of self-harm in England (3).

The probability of repeat self-harm in the next model cycle falls from 0.18 (20) to 0.11 after psychosocial assessment, based on observed experience in two of the three hospital groups covered by the MSH where the relative risk of self-harm following psychosocial assessment was 0.59 of the risk without assessment (9).

The risk of subsequent suicide in the next model cycle also draws on analysis of 13 years of self-harm hospital presentations in the MSH. This analysis indicates a very high risk of suicide in the initial six months following self-harm, but the overall rate remains 55 times greater than that of the general population after 12 months (21). Given the lack of evidence, we conservatively assumed that receipt of psychosocial assessment had no direct impact on suicide, but varied this assumption in univariate sensitivity analysis.

On average 21% of A&E attendances arrive by ambulance (22). We assumed this applied to self-harm patients, and applied a national tariff to ambulance-related costs (23). Average costs per psychosocial assessment were taken from detailed time and motion analyses previously undertaken in hospitals in Oxford and Derby that are part of the MSH (4, 20). This average cost reflects higher costs of assessments in the under-18s compared to adults. Mean treatment costs for self-harm are also taken from previous analyses of costs for admitted and non-admitted patients in Oxford and Derby hospitals in the MSH. Patterns of self-harm: poisoning only, self-injury only or a combination of the two methods, and the likelihood of hospital admission following presentation for self-harm are also based on rates seen in the MSH.

To incorporate the impact of self-harm on immediate productivity losses in the societal perspective analysis, we assumed a full day of productivity loss for each day of an inpatient stay, based on the average length of stay associated with different injuries. We assumed each productivity loss day would be 7.5 hours, valued at the national living wage rate. This is a very conservative assessment of productivity losses. Self-harm events will also mean productivity losses for individuals even if they are not admitted to hospital; while family members may also have to accompany and/or visit their relatives in hospital. No other time or out of pocket costs, such as travel costs, were considered.

Limited information is available on the quality of life impacts of self-harm, for both adults and adolescents. Previous economic evaluations of self-harm prevention have tended to use quality of life weights associated with specific mental disorders such as depression. Recently quality of life data on self-harm in 754 adolescents in England were collected using the EQ-5D-3L, an instrument widely used to elicit health related quality of life values, as part of a trial of family therapy (24). In line with this trial, we have assumed that each cycle in our model where an individual self-harms will have a quality of life weight (or utility) of 0.68. So, in a six month cycle the model assumes that there are on average 0.34 QALYs associated with experiencing self-harm and 1.36 QALYS over the model’s two-year time-horizon. This may be conservative; quality of life weights in individuals with severe mental health problems at risk of self-harm and suicide can be less than 0.5 (25). Following usual practice, death was set to a value of 0 while quality of life for those who do not self-harm is assumed to be equivalent to population norms for adolescents and young adults in England of 0.929 (26). In sensitivity analysis we vary quality of life weights down to 0.80 given the potential for enduring chronic poor mental health in some people who self-harm (27).

### Ethical standards

Although this is a modelling study drawing on data from multiple sources it should be noted that the three research sites involved in the Multicentre Study of self-harm have approvals to collect data on self-harm for their local monitoring systems of self-harm and for multicentre projects. The monitoring systems in Oxford and Derby have received their approval from local national health service research ethics committees (Oxford: South Central Berkshire REC, 08/H0607/7; Derby: Derbyshire REC, 06/Q2401/84) while self-harm monitoring in Manchester is part of a local clinical audit system ratified by the local research ethics committee (South Manchester REC). The three monitoring systems are fully compliant with the Data Protection Act (1998) and have approval under Section 251 of the National Health Service (NHS) Act (2006) to collect patient-identifiable data without explicit patient consent.

## Results

As Table 2 indicates, from an NHS perspective adherence to guidance on psychosocial assessment would potentially be cost-effective with a cost per QALY gained of £10,962 (95% uncertainty interval (UI), £15,538 – £9,219). When immediate productivity losses restricted only to inpatient time in hospital are included, the incremental cost per QALY gained falls to £9,980 (95% UI, £14,538 - £6,938). Model findings are below the notional £20,000 cost per QALY gained threshold used by NICE when making recommendations on the reimbursement of health technologies and public health interventions, although values of up to £30,000 per QALY gained can be considered appropriate where uncertainty is low (28).

**Table 2:** Incremental cost effectiveness results.

|  | Psychosocial assessment | No psychosocial assessment | Incremental cost & effect |
| --- | --- | --- | --- |
| **NHS Perspective** |  |  |  |
| Cost (95% CI) | £1223 (£1086, £1372) | £977 (£884, £1077) | £246 (£202, £295) |
| QALYs (95% CI) | 1.253 (1.085, 1.418) | 1.231 (1.072, 1.386) | 0.022 (0.013, 0.032) |
| ICER (95% UI) | £10,962 (£15,538, £9,219) |  |  |
| **Societal Perspective** |  |  |  |
| Cost (95% CI) | £1377 (£1197, £1637) | £1153 (£1009, £1411) | £224 (£189, £222) |
| QALYs (95% CI) | 1.253 (1.085, 1.418) | 1.231 (1.072, 1.386) | 0.022 (0.013, 0.032) |
| ICER (95% UI) | £9,980 (£14,538, £6,938) |  |  |

### Univariate sensitivity analysis

The sensitivity of the ICERs to individual model parameters are shown as a ‘Tornado’ diagram in Figure 1. This figure shows sensitivity of results to each parameter in a hierarchical order (i.e. parameter with greatest sensitivity at the top). The figure indicates that results are most sensitive to changes in assumptions on likelihood of repeat self-harm following psychosocial assessment. If this risk increases from the baseline value of 0.106 to 0.130 then the cost per QALY value breaches the notional £20,000 QALY threshold. If the risk of self-harm increased to 0.15, in line with the most conservative estimate of effect reported in an English study (15), then the cost per QALY would increase to £37,633. If utility values associated with self-harm were above 0.787 or if values for no self-harm fall below 0.816 then cost per QALY is above the notional threshold. Conversely, if utility values for self-harm were 20% lower than in our baseline model, then the cost per QALY gained becomes more favourable at £7,122.

**Figure 1:**
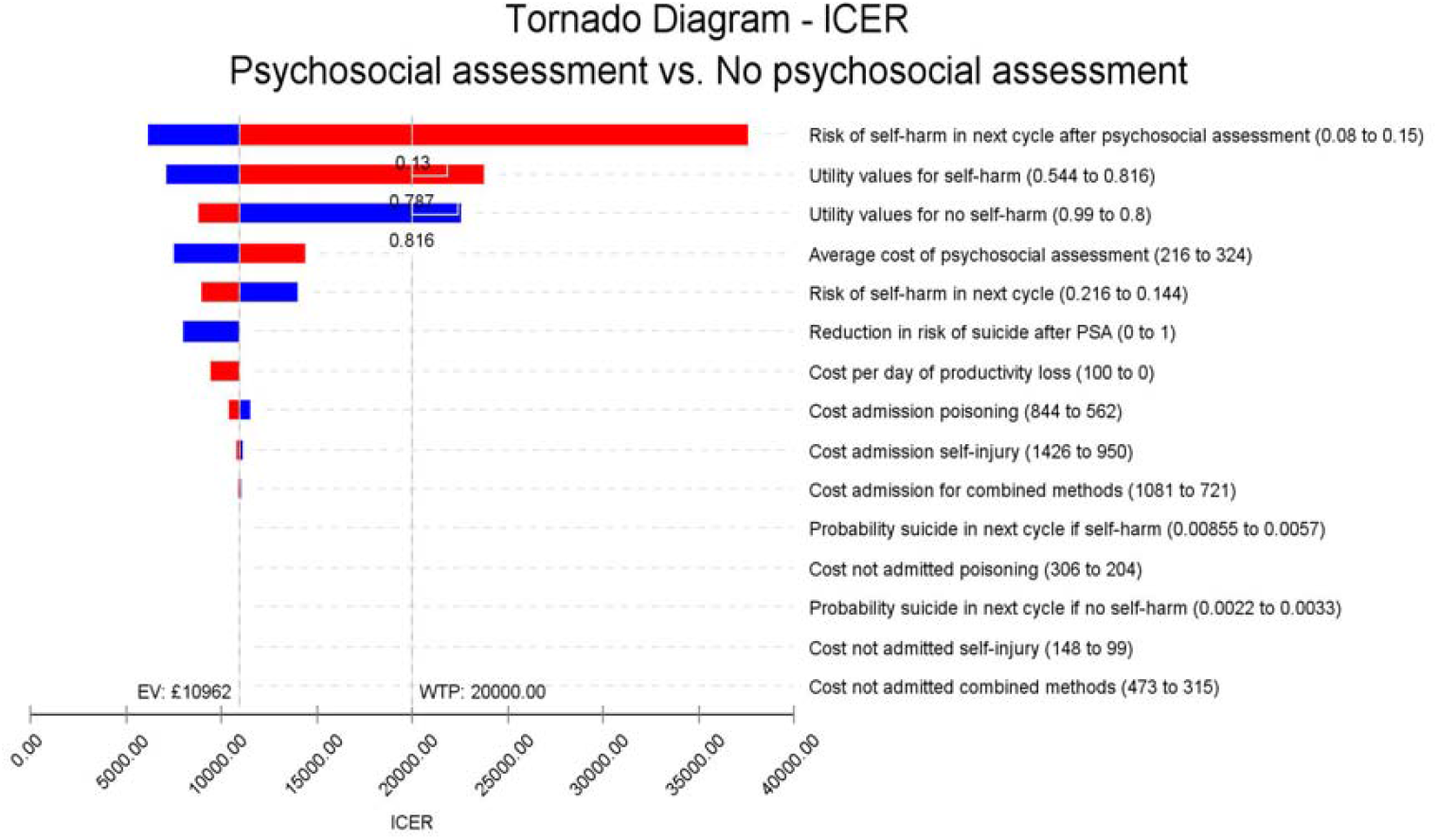
Tornado diagram. Note: The vertical line shows the mean expected incremental cost effectiveness ratio (ICER) of £10,962 per QALY gained in our base case NHS perspective scenario. Red bar segments indicate that the value of each parameter has increased, while blue segments show parameter values have fallen. Values to the right of the vertical base case scenario line indicate less favourable cost effectiveness with the cost per QALY increasing compared to the base case scenario, while those to the left indicate an improvement in cost effectiveness, with the cost per QALY gained reducing.

If the average cost of psychosocial assessment were to rise by 20% then the incremental cost per QALY would rise to £14,291, or conversely a similar fall would mean a cost per QALY of £7,484. Assumptions on likelihood of repeat self-harm also have some impact, leading to values of £13,894 and £8,883 following a 20% decrease or increase in repeat self-harm risk in any cycle.

If any direct impact between psychosocial assessment and reduced risk of suicide could be established in future, this is likely to only have a modest additional favourable impact on model findings; a 20% reduction in risk would reduce the cost per QALY to £10,195. In the unlikely event that a 100% risk reduction could be achieved the cost per QALY would fall to £7,973.

The model does not appear very sensitive to other parameters, including the costs of providing treatment for self-harm or the likelihood of inpatient admission following self-harm. If the productivity losses of all inpatient stays were included at a maximum value of £100 per day, then the cost per QALY could fall to £9,460 – compared with £9,980 when we valued each day as being worth £65.40 using the national living (minimum) wage rate.

### Probabilistic sensitivity analysis

Cost-effectiveness planes showing results of probabilistic sensitivity analysis, with 10,000 simulated ICERs from the NHS and societal perspectives are shown in Figure 2. The elliptical circles include 95% of simulations in the model. From the NHS perspective 78% of simulated pairs of costs and QALYs the use of psychosocial assessment will have an incremental cost per QALY gained below the notional £20,000 per QALY gained threshold, having higher costs than no assessment with better outcomes. Figure A3 shows the distribution of net monetary benefit (NMB) in probabilistic sensitivity analysis. This is right skewed with a mean NMB of £200 (95% CI -£189, £758).

**Figure 2:**
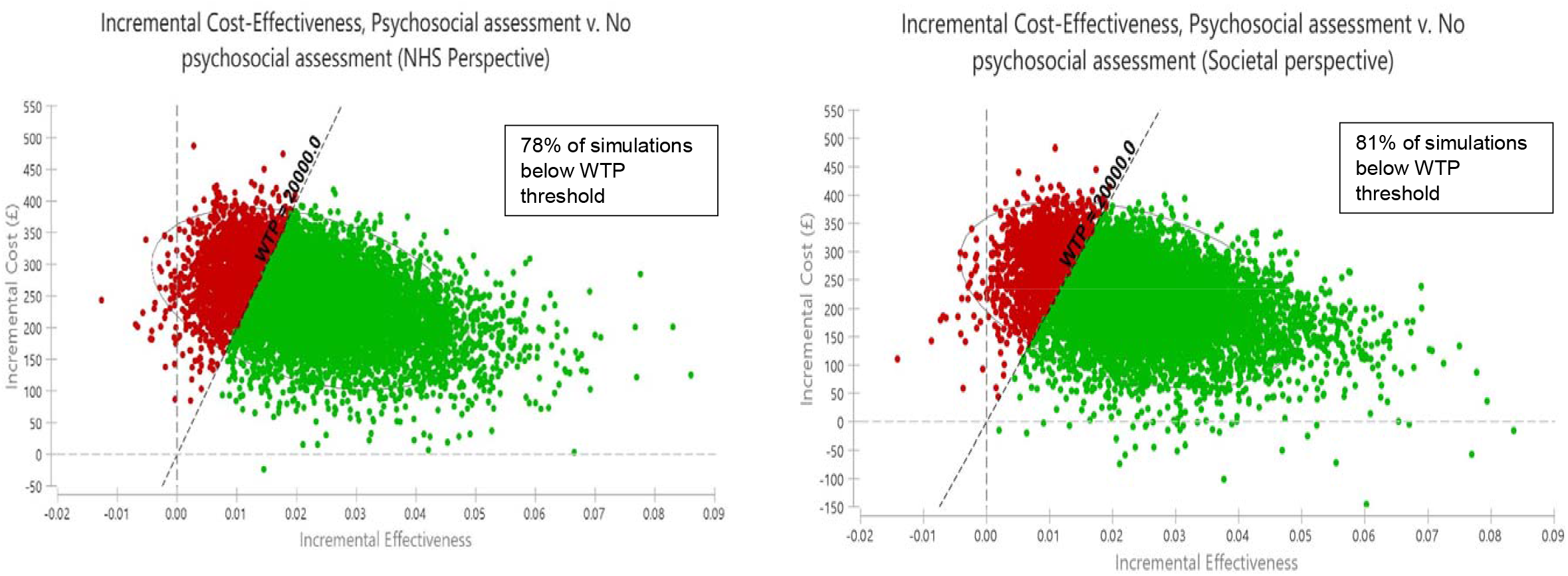
Cost effectiveness planes (NHS and societal perspectives) Note: Green dots represent simulations below the £20,000 per QALY gained cost effectiveness threshold while red dots represent simulations that are above this threshold and are not considered cost effective.

The cost-effectiveness acceptability curve in Figure A4 illustrates, for our NHS perspective model, that use of psychosocial assessment has a higher probability of being cost effective than no action if willingness to pay reaches £11,900. At a willingness to pay level of £20,000 the probability of intervention being considered cost effective is over 78%. This would rise to 91% if a higher threshold of £30,000 per QALY gained applied.

From our societal perspective there is an 81% probability of psychosocial assessment having a cost per QALY gained below £20,000 (Figure 2). Mean NMB increases to £224 (CI - £180, £801) (Figure A5). At willingness to pay levels of £11,000, psychosocial assessment has the greatest likelihood of being considered cost effective (Figure A6). The chance of being cost effective would rise to 92% if a higher threshold of £30,000 per QALY gained applied.

## Discussion

Risk of completed suicide is elevated in people who present to hospital for self-harm, being more than 50 times that of the general population and especially high in the months following hospital discharge (21). While monetary costs should not be the primary motive for improved healthcare and suicide prevention measures, past costing studies highlight very high lifetime costs of premature mortality (29, 30). Moreover, public policy decisions for similar problems, such as injury prevention and road safety, commonly take the costs and benefits of measures to reduce avoidable deaths into account. Health policy makers have also examined the economic case for investment in interventions to prevent suicide and self-harm, with the National Institute for Health and Care Excellence (NICE) in England commissioning economic analysis as part of the development of national guidelines on suicide prevention (31, 32).

Current national clinical guidance on management of self-harm recommends psychosocial assessment should be offered to all individuals each time they present at hospital after self-harm (2, 6). However, there is evidence of great variation in receipt of assessment in English hospitals (7, 8, 33). While studies have not been designed to evaluate the effectiveness of psychosocial assessment alone, some studies have suggested that assessment may be associated with reduced future risk of hospital-presenting self-harm. Potentially this may be partly due to receipt of more appropriate subsequent primary and community care, as well as the therapeutic benefits of the psychosocial assessment process itself.

We believe this is the first economic evaluation specifically on benefits of increasing use of psychosocial assessment for all hospital presenting self-harm. Our analysis suggests there is an economic case for adherence to NICE guidance, with a 78% chance from a health service perspective of cost per QALY gained being under the notional £20,000 threshold considered cost-effective by decision makers in England. Likelihood of psychosocial assessment being cost-effective was even higher when taking the societal perspective, even without incorporating impacts of reduced self-harm repetition on family members or productivity losses associated with completed suicide. Our estimates are conservative as our analysis only covers two-years; cost-effectiveness would be greater if benefits persist over a longer time-period.

Our findings are consistent with analysis of immediate impact of extending hours of a liaison psychiatry service at another English hospital, with a view to increasing the number of individuals who received a psychosocial assessment (34). After liaison psychiatry service hours were extended the proportion of individuals attending A&E following self-harm who received psychosocial assessment increased from 57% to 68%, and the additional costs of liaison psychiatry were partly offset by a 14% reduction in hospital costs related to the management of self-harm. That study also observed a plausible 20% reduction in the future risk of hospital presenting self-harm, but noted that it was not sufficiently powered to detect any significant effect on self-harm rates. Analysis in Scotland of a very brief hospital administered psychological therapeutic intervention immediately after self-harm was also shown likely to be cost effective, particularly for individuals with a history of self-harm (35).

Strengths of our modelling analysis include reliance on a large dataset across multiple hospitals where robust methods are used to identify all self-harm presentations regardless of whether these result in hospital admission (3). We have also made use of detailed costing analyses using this dataset to estimate costs associated with psychosocial assessment and immediate treatment (20). Conversely, because our model draws on data on self-harm presentations from a limited number of locations in England, these may not be representative of patterns of self-harm nationally, but our model was sensitive neither to changes in patterns of self-harm nor in rates of inpatient admission.

We also recognise that there appears, understandably, to have been no specific randomised controlled study looking solely at the role of psychosocial assessment in directly reducing future risk of self-harm. The magnitude of any therapeutic impact of psychosocial assessment on relative risk of repeat self-harm may vary considerably, depending, for example, on its quality. Our baseline probability of repeat self-harm after psychosocial assessment of 0.11 draws on data from two of the three hospital groups covered by the MSH that had a relative risk reduction of 0.59 following psychosocial assessment (9). Other studies have reported more conservative impacts. One study reported a relative risk of 0.7 for repeat self-harm, but this was based on experience in just one of the MSH sites, making its generalisability more limited (16). Another English study using cohort data from three hospitals reported a relative risk of 0.82 using instrumental variables (IV) methodology, but cautioned the estimate could be biased because of limitations in the application of the IV methodology (15). A previous modelling analysis, looking at ‘risk scales’ as part of a hospital assessment process, assumed a relative risk of 0.8 for individuals receiving a psychosocial assessment (17). In our sensitivity analysis the probability of repeat self-harm following psychosocial assessment needs to be 0.13 or lower (equivalent to a relative risk of 0.73 for repeat self-harm) for the intervention to be considered cost-effective from a health system perspective at a cost per QALY threshold of £20,000.

More information is also needed on the quality of the psychosocial assessment process, as well as content, skills and formulation; this has not been a feature of previous analyses and is very likely to have a bearing on effectiveness (36). It should also be noted that in our model we have held the likelihood of each psychosocial assessment reducing the risk of future self-harm constant; however, receipt of multiple psychosocial assessments may improve the level of risk reduction (16). This merits further investigation.

Our sensitivity analysis indicates the model is sensitive to assumptions about quality of life. Few studies have elicited quality of life values for hospital presenting self-harm; we were able to use EQ-5D-3L quality of life estimates collected as part of an English trial of family therapy following self-harm, but this study only examined quality of life for adolescents (24). We identified another English study where quality of life scores following self-harm were lower, but that study only included adult psychiatric inpatients (25). Conservatively, we have also not included impacts on quality of life of the family and friends of individuals who self-harm, although these impacts are not normally included in the NICE decision-making process.

Our modelling analysis only considered the impact on immediate assessment and treatment costs in hospital; we were unable to look at impacts on longer-term primary and specialist community mental health team use. Success in reducing self-harm and completed suicide will partly depend on the longer-term care and support individuals receive after leaving hospital. Psychosocial assessment may increase the likelihood that individuals make use of appropriate services, and these costs have not been included. However, some of these costs are likely to be for treatment of physical and mental health problems unrelated to self-harm behaviour. The extent to which these knock-on costs explicitly link with self-harm needs to be investigated.

This need for future work to look at long-term health service utilisation has also been highlighted in other comparable studies (34). One previous English study looking at long-term health and social care service utilisation of a small number of individuals following an initial self-harm event suggests that continuity of care may be relatively modest; average cumulative costs per individual over seven years were £3,991 (2020 prices) (37). However, this study also reported much higher costs for individuals with multiple repeat events. If future risk of self-harm is reduced through psychosocial assessment, then we may have omitted benefits of future avoided costs through better self-harm management and underestimated the economic case for psychosocial assessment. Long term impacts beyond the health sector, such as lost lifetime earnings linked to self-harm, might also be assessed (38).

## Conclusions

Psychosocial assessment as implemented in the English NHS is likely to be cost-effective. This first evidence could support efforts to increase adherence to NICE guidelines. However, further evidence about the precise impact of psychosocial assessment on self-harm repetition, quality of life benefits of self-harm avoidance and costs to individuals affected by self-harm and their families beyond immediate hospital stays, is still needed. Given variable adherence to guidelines in England it is also imperative to explore the cost effectiveness of mechanisms that could change health care professional practice to better improve adherence.

## Data Availability

Individual patient-level from the Multicentre Study of self-harm will not be available due to confidentiality and data-sharing agreements in place.

## Author contribution

DMD, A-LP, AT and KH conceived the idea of the study. DMD and A-LP developed the models, drafted the manuscript, led the analyses and interpreted the results alongside AT, FB, DC, CC, GG, NK, JN, KW and KH. All authors made revisions to earlier drafts and approved the final manuscript.

## Data availability

Data availability is not applicable as no new data were created or analysed in this study. The modelling analysis did make use of data from the Multicentre Study of self-harm, however individual patient-level from the Multicentre Study of self-harm will not be available due to confidentiality and data-sharing agreements in place.

