## Supplementary Figures for "Cost-effectiveness of psychosocial assessment for individuals who present to hospital following self-harm in England: a model-based retrospective analysis"

**Supplementary Appendix: Additional Figures and Table**

**Figure A1: Overview of 3 state Markov model**

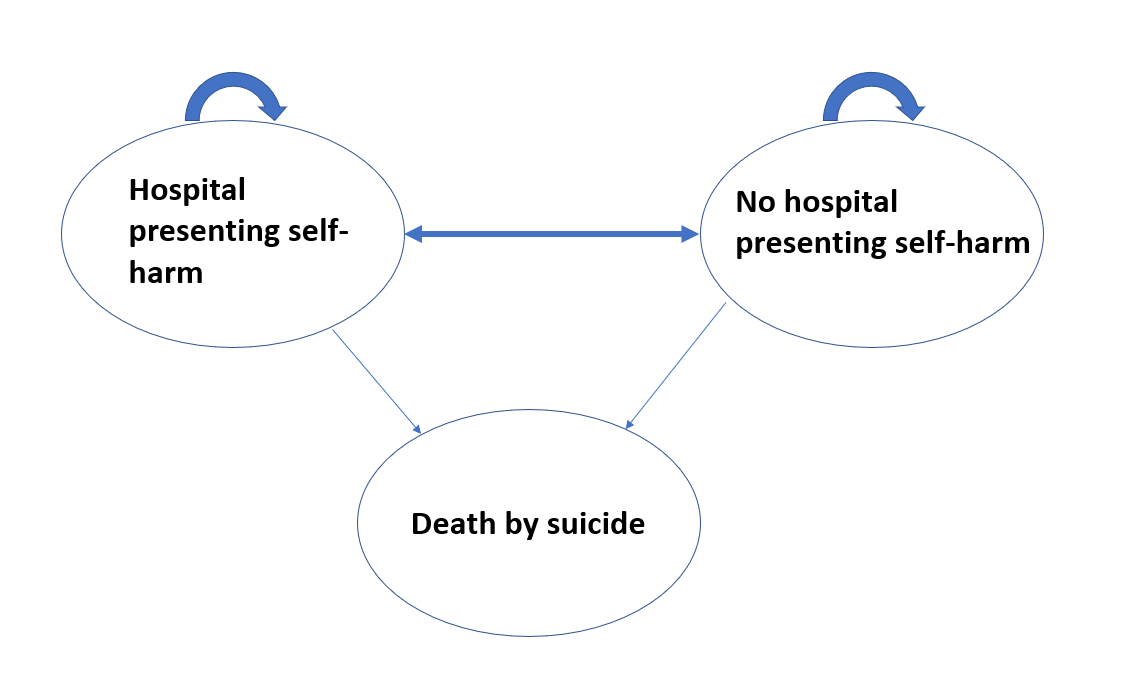

**Figure A2: Detailed excerpt of model structure**

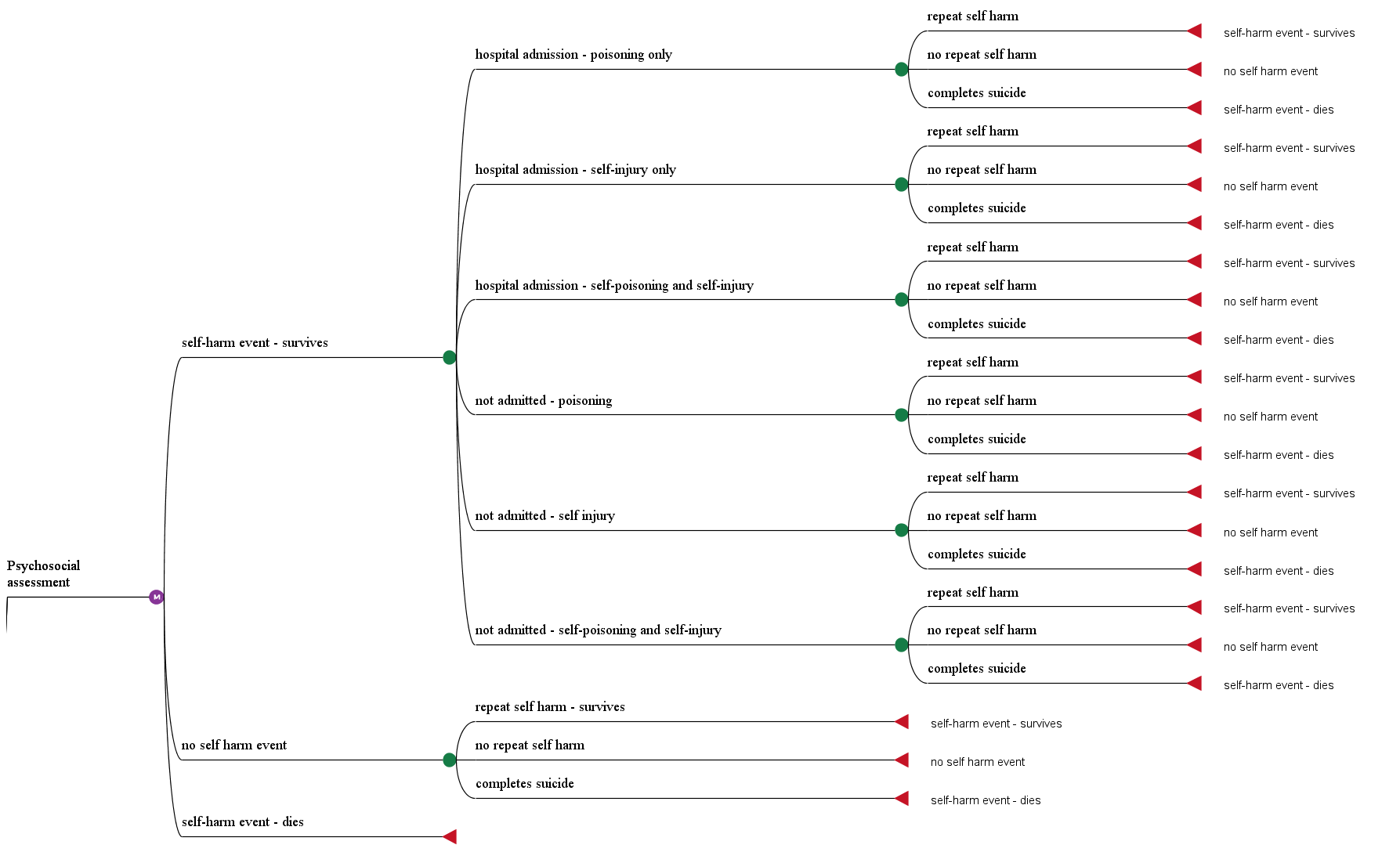

Note: Model structure is the same for the no intervention pathway (not shown)

**Figure A3: Net monetary benefit probability distribution (NHS perspective 10,000 bootstraps)**

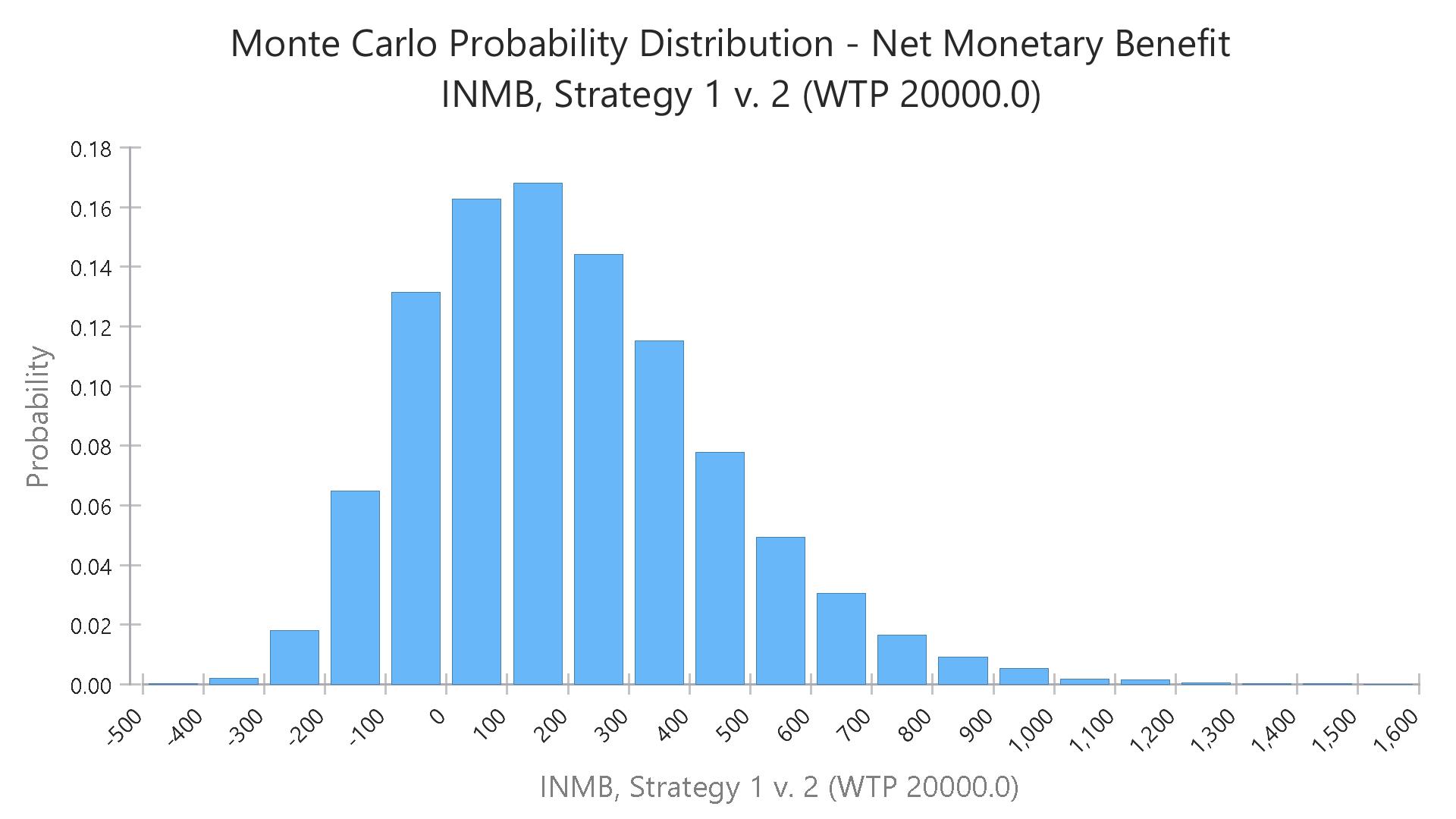

Mean NMB: £200

(95% CI -£189, £758)

**Figure A4: Cost effectiveness acceptability curve (NHS perspective)**

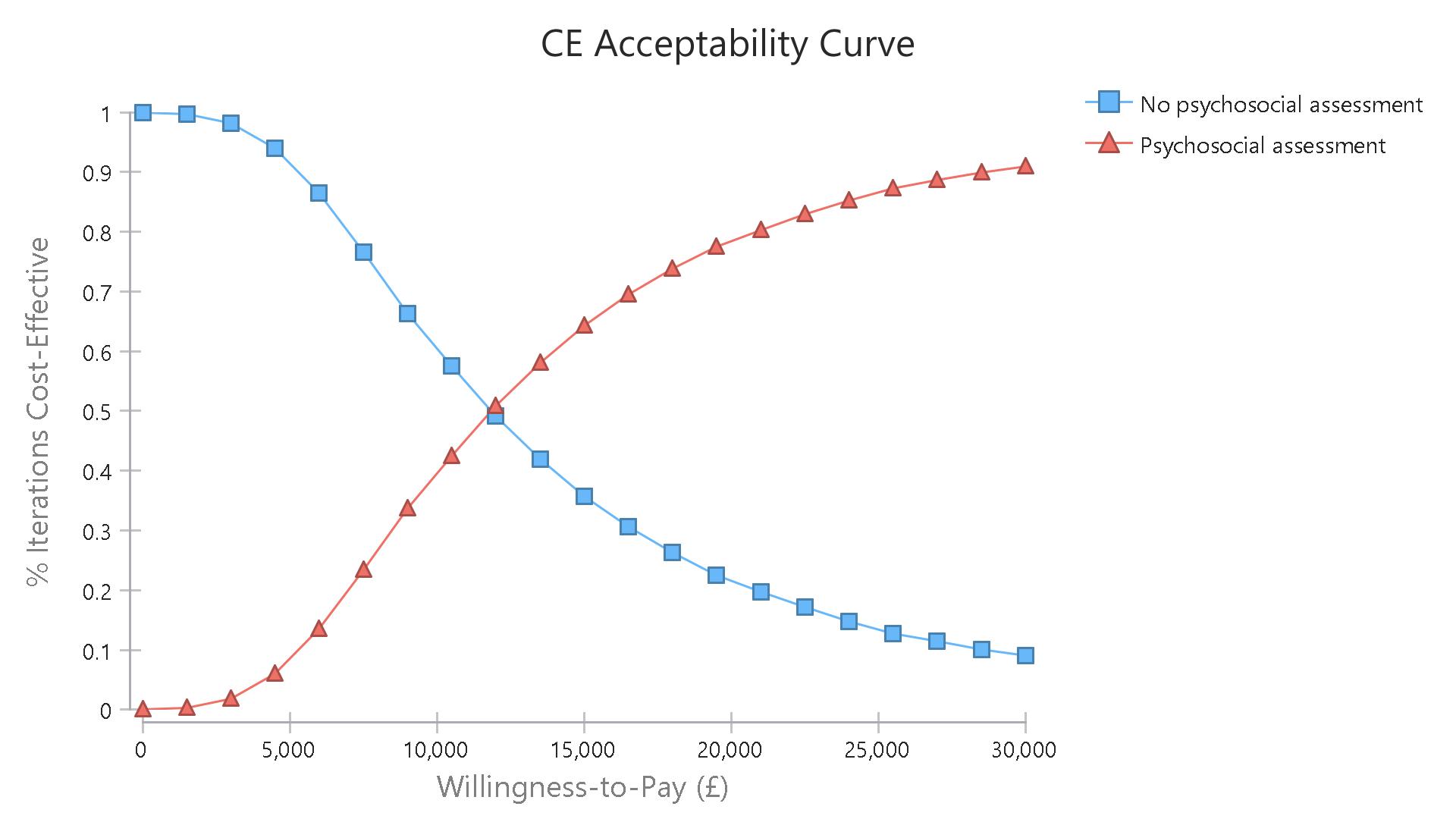

£11,900

**Figure A5: Net monetary benefit probability distribution (Societal perspective 10,000 bootstraps)**

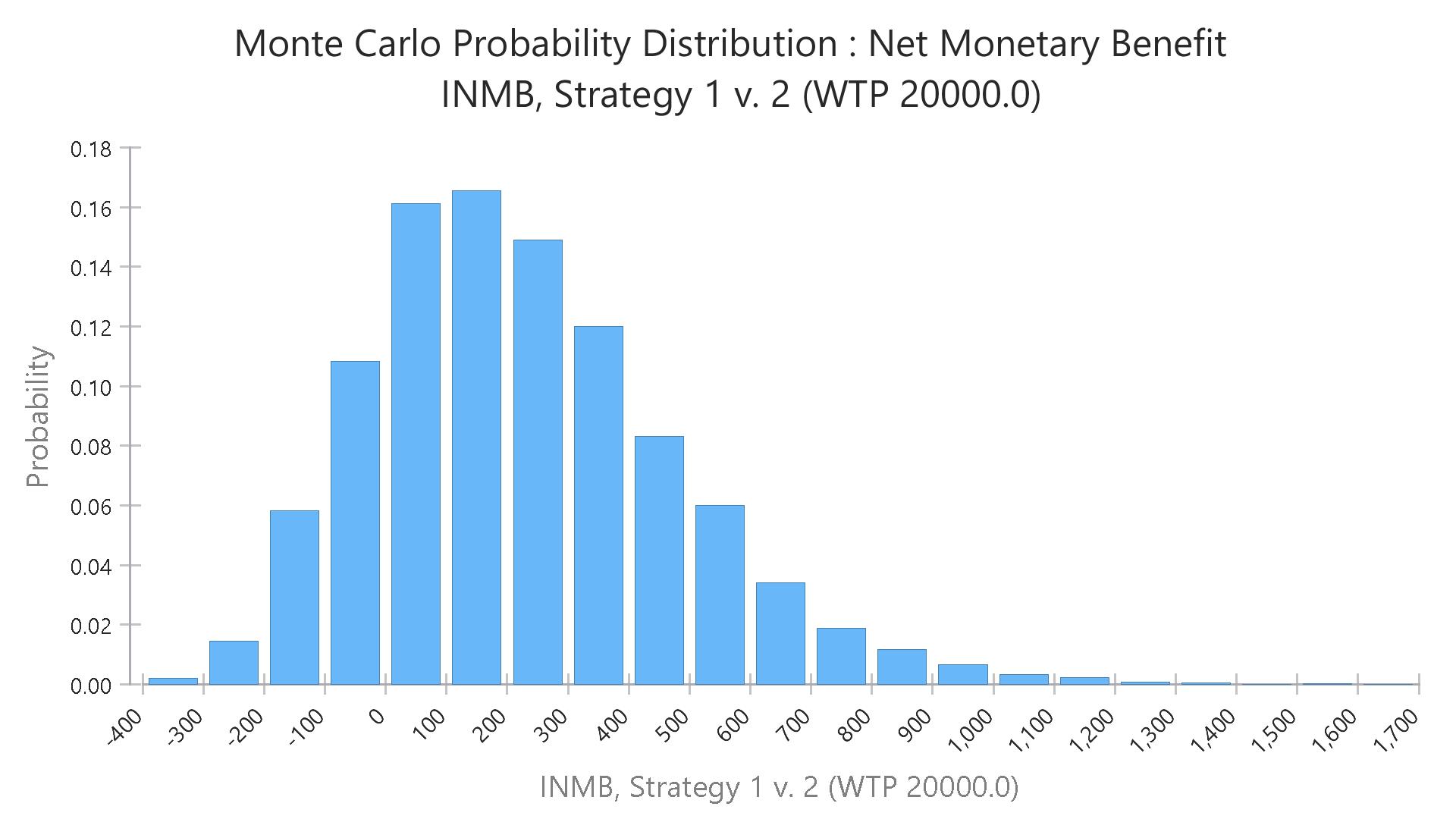

Mean NMB: £224

(95% CI -£180, £801)

**Figure A6: Cost effectiveness acceptability curve (societal perspective)**

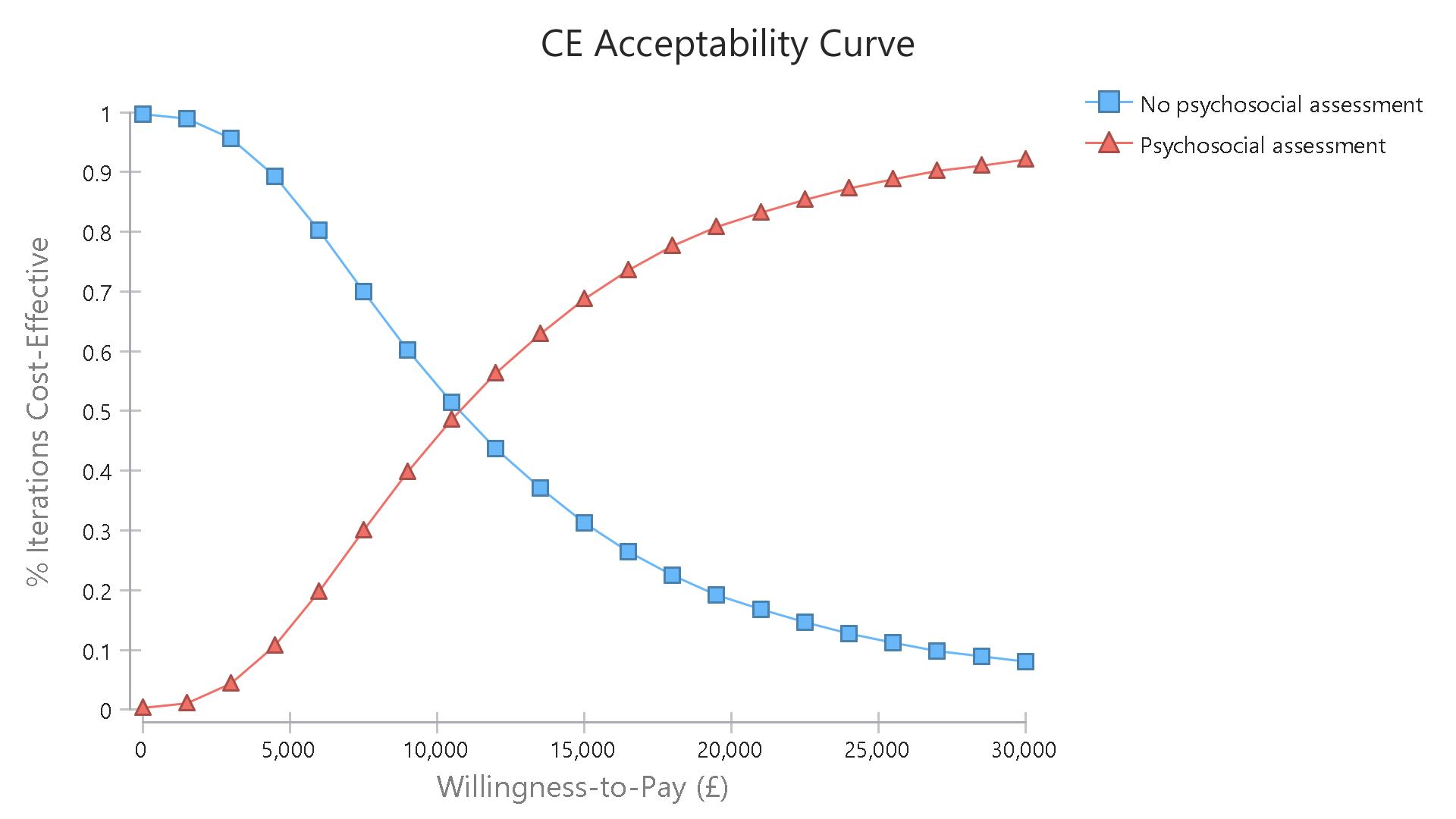

£11,000
